# Human blood DNA methylation patterns reflect individual lifestyle

**DOI:** 10.1101/2021.12.16.21267895

**Authors:** Ireen Klemp, Anne Hoffmann, Luise Müller, Tobias Hagemann, Kathrin Horn, Kerstin Rohde-Zimmermann, Anke Tönjes, Joachim Thiery, Markus Löffler, Ralph Burkhardt, Yvonne Böttcher, Michael Stumvoll, Matthias Blüher, Knut Krohn, Markus Scholz, Ronny Baber, Paul W Franks, Peter Kovacs, Maria Keller

**Affiliations:** Medical Department III – Endocrinology, Nephrology, Rheumatology, University of Leipzig Medical Center, Leipzig 04103, Germany; Helmholtz Institute for Metabolic, Obesity and Vascular Research (HI-MAG) of the Helmholtz Center Munich at the University of Leipzig and University Hospital Leipzig, Leipzig, 04103, Germany; Institute for Medical Informatics, Statistics and Epidemiology, University of Leipzig, 04103 Leipzig, Germany; Medical Faculty, University of Kiel, 24118 Kiel, Germany; Institute of Clinical Chemistry and Laboratory Medicine, University Hospital Regensburg, 93053 Regensburg, Germany; Department of Clinical Molecular Biology, Institute of Clinical Medicine, University of Oslo, 0316 Oslo, Norway; Medical Division, Akershus University Hospital, 1478 Lørenskog, Norway; Deutsches Zentrum für Diabetesforschung, 85764 Neuherberg, Germany; Medical Faculty, University of Leipzig, 04103 Leipzig, Germany; LIFE Leipzig Research Center for Civilization Diseases, University of Leipzig, 04103 Leipzig, Germany; Genetic and Molecular Epidemiology Unit, Department of Clinical Sciences, Lund University, Skåne University Hospital, 21428 Malmö, Sweden

**Keywords:** lifestyle score, diet, physical activity, smoking, alcohol, epigenetics, DNA methylation, epigenetic clock

## Abstract

Obesity is driven by modifiable lifestyle factors whose effects may be mediated by epigenetics. Therefore, we investigated lifestyle effects (diet, physical activity, smoking and alcohol) on blood DNA methylation in participants of the LIFE-Adult study, a well-characterized population-based cohort from Germany. Fifty subjects with an extremely healthy and 50 with an extremely unhealthy lifestyle were selected for genome-wide DNA methylation analysis in blood samples. Whereas obesity was only marginally related to variability in DNA methylation pattern, comparisons between lifestyle categories resulted in 145 Differentially Methylated Positions (DMPs) and 4682 Differentially Methylated Regions (DMRs) annotated to 4426 unique genes. Intersection analysis showed that diet, physical activity, smoking and alcohol intake are equally contributing to the observed differences, which particularly affects pathways related to glutamatergic synapse and axon guidance. DNA methylation patterns help discriminate individuals with a healthy vs. unhealthy lifestyle, which may mask subtle methylation differences derived from obesity.

## Objective

Obesity is well recognized as a multifactorial disease in most modern societies, with not only individuals’ genetic background contributing to the disease burden, but also with a crucial role of lifestyle and environment, strongly influencing epigenetic mechanisms controlling metabolic processes ^1^. However, common lifestyle intervention regimes vary greatly in structure and length and, thus, in their individual success on weight reduction [reviewed in Aronica et al. ^2^]. Observed direct effects e.g. on DNA methylation patterns after short-term lifestyle interventions are often marginal ^2^, which might be due to their short duration and low intensity ^3^. Recent studies demonstrated that successful short term weight loss interventions may reduce methylation age (mAge) to the chronological age level ^4^. Furthermore, DNA methylation patterns may predict the success of lifestyle-induced weight loss ^5–7^.

Comprehensive studies investigating the underlying interaction between genetics, epigenetics and especially lifestyle are currently lacking. Therefore, we I) analyzed and compared the human blood DNA methylation patterns between subjects living a healthy and those living an unhealthy lifestyle. II) We further compared obese and non-obese subjects to identify DNA methylation patterns, which are related to an obese phenotype despite a healthy lifestyle, or potentially associated with a healthy (lean) phenotype in an allegedly unhealthy obesogenic environment. III) We elucidated lifestyle specific effects on the epigenetic clock. IV) Finally, we investigated the role of genetic variants cis to the identified target regions by meQTL (methylation quantitative trait loci) analyses and addressed potential consequences of these changes on blood transcriptome by matrix-eQTMs (expression quantitative trait methylations).

## Research Design and Methods

### Study population

The present analyses included participants of the LIFE-Adult study, a population-based cohort focusing on lifestyle diseases ^8^. The cohort is comprised of ∼10,000 adult subjects aged from 18 to 80 years (mean± standard deviations (SD): age = 57.4±12.5 years, BMI = 27.3±4.9 kg/m²) from the region of Leipzig, Germany. All participants underwent an extensive phenotyping including anthropometric measurements, social- and lifestyle-behavior questionnaires and blood parameters. For most subjects, EDTA blood samples are available ^8^. All participants gave written informed consent to participate in the study and procedures were approved by the University of Leipzig’s ethics committee (registration number: 263-2009-14122009) and conducted according to the Declaration of Helsinki. Study participation, assessments and interviews were supervised and carried out by trained staff and under supervised standard operation procedures ^8^.

### Lifestyle Score

We created a Lifestyle Score (LS) as sum of four different sub scores: diet, physical activity, alcohol consumption and smoking ^9^. To calculate the scores, we included data from four self-reported questionnaires: I) a German version of the Food Frequency Questionnaire (FFQ) ^10^, II) the Short-Form International Physical Activity Questionnaire (SF-IPAQ) ^11^, III) a questionnaire about smoking status and quantity, as well as IV) about daily alcohol consumption and frequency. The final LS ranged from 3 to 66 (mean±SD: 27.19±11.2), with low and high LS values representing a healthy and unhealthy lifestyle, respectively. Detailed description of the individual scoring as well as explanation of each sub-score can be found in Supplemental Material and Supplemental Table 1. Subjects with any missing questionnaire item were completely excluded from further analyses to avoid potential effects caused by general non-compliance of those subjects. Similarly, participants with pre-existing diabetic conditions (HbA1c≥6.5%) ^12^ or missing BMI measures were also excluded from subsequent analyses. A total of 4107 subjects (mean±SDs: Age=56±13 years, BMI=27.0±4.7 kg/m², LS=27.19±11.02) passed all criteria (Table 1).

**Table 1.** -Study Characteristics: Phenotypic data are described for all in the Lifestyle Score (LS) analysis included LIFE-adult subjects as well as for the healthy (≤5^th^ percentile) and unhealthy (≥95^th^ percentile) living extreme subgroups as mean±SD (incl. discovery and validation cohort; detailed in Supplemental Table 2). *P*-values show significant differences between healthy and unhealthy subgroups.

|  | **LIFE cohort** | **Healthy Lifestyle (LS≤5<sup>th</sup> pct.)** | **Unhealthy Lifestyle (LS≥95<sup>th</sup> pct.)** | **P-values (LS 5th vs. 95th pct.)** |
| --- | --- | --- | --- | --- |
| **N (total number)** | 4107 | 216 | 207 |  |
| **Gender (N: female/male)** | 2109/1998 | 160/56 | 66/141 |  |
| **Age (years)** | 55.9 ± 12.8 | 60.14 ± 12.76 | 54.03 ± 10.47 | <0.001 |
| **BMI (kg/m<sup>2</sup>)** | 27.03 ± 4.65 | 27.11 ± 4.74 | 27.31 ± 4.69 |  |
| **BMI category (N: lean/obese/overweight)** | 1488/1659/940 | 73/95/48 | 67/93/46 |  |
| **Waist circumference (cm)** | 95.71 ± 13.02 | 93.46 ± 12.18 | 99.52 ± 13.59 | <0.001 |
| **Waist-to-Hip Ratio** | 0.93 ± 0.09 | 0.9 ± 0.08 | 0.97 ± 0.08 | <0.001 |
| **Fasting plasma glucose (mmol/l)** | 5.56 ± 0.79 | 5.63 ± 0.79 | 5.61 ± 0.72 |  |
| **Fasting plasma insulin (pmol/l)** | 63.33 ± 42.82 | 58.53 ± 36.99 | 66.66 ± 43.77 |  |
| **Plasma Low Density Lipoprotein (mmol/l)** | 3.5 ± 0.95 | 3.5 ± 0.93 | 3.63 ± 0.97 |  |
| **Plasma High Density Lipoprotein (mmol/l)** | 1.62 ± 0.46 | 1.79 ± 0.45 | 1.44 ± 0.42 | <0.001 |
| **Plasma Apolipoprotein A1 (g/l)** | 1.67 ± 0.3 | 1.76 ± 0.27 | 1.6 ± 0.3 | <0.001 |
| **Plasma Triglycerides (mmol/l)** | 1.38 ± 1.1 | 1.17 ± 0.58 | 1.68 ± 1.03 | <0.001 |
| **Diet Score** | 12.39 ± 3.22 | 8.69 ± 1.94 | 15.39 ± 3.08 | <0.001 |
| **Physical activity Score** | 7.55 ± 6.47 | 0.58 ± 1.6 | 17.25 ± 3.8 | <0.001 |
| **Smoking Score** | 5.91 ± 6.70 | 0.09 ± 0.68 | 16.81 ± 3.58 | <0.001 |
| **Alcohol Score** | 1.34 ± 2.22 | 0.09 ± 0.68 | 2.97 ± 2.46 | <0.001 |
| **LS** | 27.19 ± 11.02 | 9.45 ± 1.54 | 52.42 ± 4.07 | <0.001 |

**Table 2.**
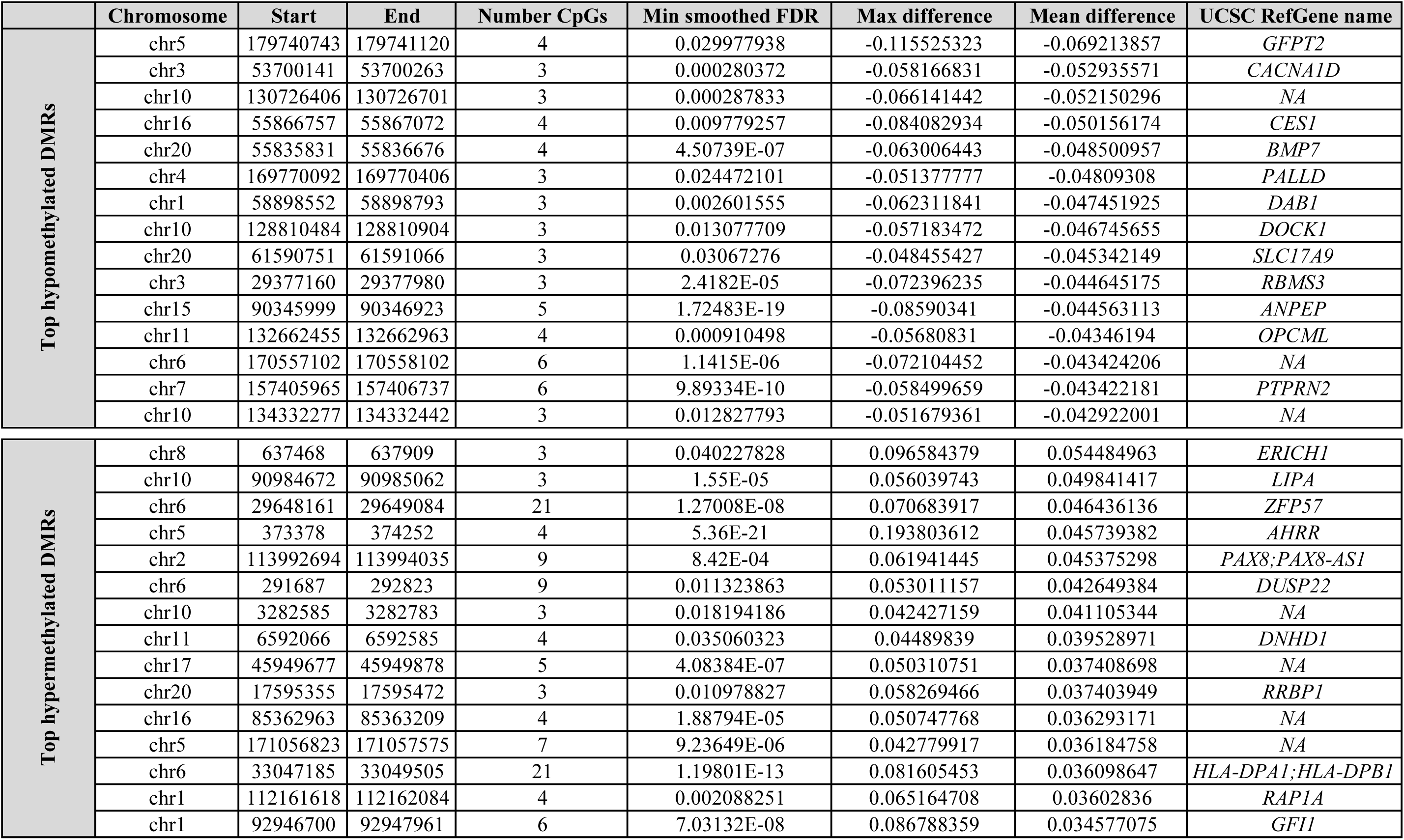
-Top lifestyle-specific Differentially Methylated Regions (DMRs): Included are the top 15 significantly hyper- and hypo methylated DMRs (min. smoothed FDR<5%) between healthy and unhealthy living subjects.

### Subset for genome-wide methylation and validation measurements

Based on the LS calculation we stratified the cohort into two groups reporting the most healthy and unhealthy lifestyles, by selecting the lowest and highest five percent (5^th^ percentile LS≤11; 95^th^ percentile LS≥48). Within these groups (*N*=234), we found 140 subjects with and 94 subjects without obesity according to BMI criteria ^13^. Based on this and an equal age range (Supplemental Figure 1b) we further selected 25 subjects without (BMI<25 kg/m²) and 25 subjects with obesity (BMI>30 kg/m²) amongst each subgroup (5^th^ vs. 95^th^ percentile) (*N* total=100) for the genome-wide methylation discovery cohort and included all (with and without obesity) subjects with sufficient available DNA (*N*=213) for validation analysis. The discovery cohort comprised 44 males and 56 females with a mean BMI of 28.6±6.5 kg/m². The validation cohort (*N*=213; 5^th^/95^th^ percentile=100/113; lean/obese=131/82) included 84 males and 129 females with a mean BMI of 27.1±6.2 kg/m² (Supplemental Table 2).

### Sample preparation

All samples were isolated, stored and maintained at the Leipzig Medical Biobank ^14^, according to standard protocols. Briefly, blood samples were taken after an overnight fast (mean fasting duration 12.7±1.7h) during the individuals’ study visit and stored at 4-8 °C until DNA isolation (within 48h after blood withdrawal) on the Autopure LS platform (Qiagen, Germany) using chemistry by Qiagen and Stratec Molecular (Stratec, Germany). Genomic DNA samples have been stored at -80°C prior integrity control using gel-electrophoresis and concentration measurements of double stranded DNA using Quant-iT PicoGreen dsDNA (Invitrogen, ThermoFisher Scientific, Germany) and Quantus (Promega, Germany) technologies.

### Genome-wide DNA methylation analysis

500ng of genomic DNA was taken for bisulfite conversion using EZ DNA Methylation Gold Kit (Zymo Research, Netherlands). After quality control, amplification, and hybridization on Illumina HumanMethylation850 Bead Chips (Illumina, Inc., San Diego, CA, U.S.A) the Illumina iScan array scanner was used to quantify genome-wide DNA methylation levels at 850K CpG sites per sample on single-nucleotide resolution.

Raw data was first quality controlled using the QC report of the minfi R package (v1.38.0) ^15, 16^. Two samples, which do not pass the badSampleCutoff of 10.5, were excluded during normalization steps. Beta densities and control probes were within predicted specifications. Probes which did not passed detection *P*-value (*P*_detect_=0.01) in more than 1% of all 98 samples were excluded from the analysis (17,375 probes). Cross-reactive probes (38,924 probes) ^17^ and probes containing known single nucleotide polymorphisms (SNPs) (29,383 probes) at the CpG site were also filtered out applying the maxprobes (v0.0.2) and minfi R packages, respectively. In addition, probes on sex chromosomes were removed from the analysis subset (19,627 probes), as sex represents a larger source of variation in our methylation data. In total 760,550 probes remained for the analysis. ß-value generation and quantile normalization was computed using the minfi R package ^15, 16^ and adjusted for the sex specific batch effects (see Supplemental Figure 1a). Further, we analyzed the cell type composition using the Houseman approach ^18–20^ adapted to EPIC arrays by Salas et al. ^20^. Possible differences in cell type composition were (see Supplemental Figure 2) analyzed using Wilcoxon tests in R. We corrected ß-values for cell type composition in an attempt to reduce noise ^20^, although none of the cell type populations differed strongly between the subgroups (low and high LS) (Supplemental Figure 2a) and the low and high LS subgroups in individuals without and with obesity (Supplemental Figure 2b).

Differential methylation analyses were performed between subjects with extreme healthy and unhealthy lifestyle (low vs. high LS/ 5^th^ *vs.* 95^th^ percentile) as well as between participants without (BMI<25 kg/m²) and with obesity (BM>30 kg/m²) within each lifestyle subgroup. The established R package limma (v3.48.3) was used for identifying differentially methylated CpG sites ^21^. As the arrays were run on two lanes, the different array slides were included as covariates in the analyses for DMPs and DMRs (Supplemental Figure 3). DMPs with an adj. *P*-value<5% were defined as differentially methylated. Differentially methylated regions (DMRs) were extracted applying DMRcate (v2.6.0) ^22^ which uses a Gaussian kernel smoothing to find patterns of differential methylations independent of genomic annotation. Only DMRs with more than two CpG sites were reported. DMRs with a min. smoothed FDR<5% were defined as differentially methylated. DMRs with a mean methylation difference >±2% were further annotated to CpG islands (CpG shores, CpG shelves and inter-CGI) and gene context related regions (promoters, 5’UTRs, exons, introns, 3’UTRs and intergenic). Genomic annotation were performed using the annotatr R package (v1.18.1) ^23^, with respect to multiple annotations. To elucidate putative drivers of blood DNA methylation, separate analysis with individual covariates (smoking, diet, physical activity, alcohol, BMI and age) were performed. Intersection analysis for covariate specific effects on lifestyle DMRs was performed using the UpsetR (v1.4.0) package ^24^.

### Methylation age- and telomere lengths-clocks

DNA methylation age (DNAmAge), corresponding DNAmAge acceleration differences according to Horvath’s clock (I and II), Levine’s clock and the telomere lengths clock were estimated using the R package methylclock v. 0.7.5 ^25^.

### KEGG-pathway overrepresentation

Candidate genes identified by significant DMRs (min. smoothed FDR<5%) characterizing lifestyle specific methylation differences (healthy vs. unhealthy living subjects) and differences between subjects without and with obesity were taken forward for a KEGG pathway over-representation test using clusterProfiler::enrichKEGG (v.3.18.1) ^26^. Enrichment *P*-values were adjusted using Benjamini-Hochberg correction and FDR<5% was considered as statistically significant.

### Validation of candidate CpGs

We selected two top candidate DMPs (Table 3, Supplemental Table 8) from our discovery cohort (high LS *vs*. low LS) for additional validation using bisulfite sequencing. Briefly, 300ng of genomic DNA were bisulfite converted using EpiTect Fast DNA Bisulfite Kit (Qiagen, Germany). After a whole genomic amplification using the EpiTect Whole Bisulfitome Kit (Qiagen, Germany) candidate regions were amplified and sequenced using the PyroMark Q24 platform and self-designed assays for *Retinoic Acid Receptor Alpha (RARA)* and *F2R Like Thrombin Or Trypsin Receptor 3* (*F2RL3)* candidate DMPs (Qiagen, Germany). Primer sequences are shown in Supplemental Table 3. All analyses were performed in duplicates including two non-template controls per sequence run.

**Table 3.**
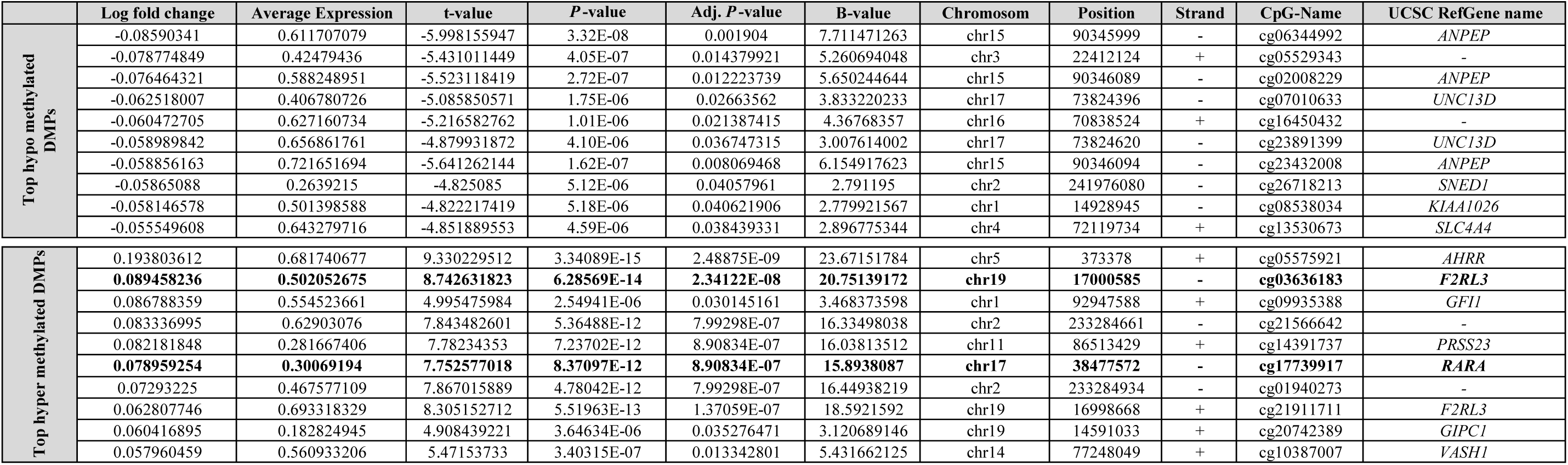
-Top lifestyle-specific Differentially Methylated Positions (DMPs): Included are the top 10 significantly hyper- and hypo methylated DMPs (adj. *P*-value<0.0**5**) between healthy and unhealthy living subjects. Our selected candidates for the bisulfite validation are highlighted.

### Transcriptome data

Transcriptome data was available from Illumina HT-12 v4 Expression BeadChips (Illumina, San Diego, CA, USA) using whole blood RNA samples from the LIFE-Adult cohort as described elsewhere ^8, 27^. Data processing was performed using R/Bioconductor after extraction of all 47,231 gene-expression probes using the Illumina GenomeStudio without background correction. Further, expression values were log2-transformed, quantile-normalized ^28, 29^ and batch effect correction was performed using an empirical Bayes method ^30^. Probes were excluded when expressed in less than 5% of the (subgroup-specific) samples (detected by Illumina GenomeStudio), still being associated with batch effects after Bonferroni-correction or not mapping to a gene accordingly ^31^ (accessed on 2019-04-04). Additionally, gene probes without available annotation and genes on X and Y chromosomes were removed in order to effects introduced by sex. In summary 20,114 valid gene-expression probes were identified corresponding to 14,687 single genes in the human genome (hg19). A three step procedure was used to remove bad quality samples: I) first, the number of detected gene-expression probes of a sample was required to be within ± 3 interquartile ranges (IQR) from the median, II), the Mahalanobis distance of several quality characteristics of each sample (signal of AmbionTM ERCC Spike-In control probes, signal of biotin-control-probes, signal of low-concentration control probes, signal of medium-concentration control probes, signal of mismatch control probes, signal of negative control probes and signal of perfect-match control probes) ^32^ had to be within median + 3 x IQR and III), Euclidean distances of expression values as described ^29^ had to be within 4 x IQR from the median. Overall, of the assayed 3,526 samples, 107 samples were excluded for quality reasons.

### Genotype data

For genotypes, 7,838 participants of LIFE-Adult were genotyped using the genome-wide SNP array Affymetrix Axiom CEU1 and the software Affymetrix Power Tools (version 1.20.6). Quality control (QC) of the genotyped data was performed following Affymetrix’s data analysis guide ^33^ as previously described ^34^. Quality control according to Affymetrix’s data analysis guide included dish-QC (<0.82), sample call rate (<97%), sex-mismatch, ambiguous relatedness (e.g. sample mix-up) and abnormalities of XY intensity plots (e.g. XXY samples filtered for gonosomal analyses). Genetic heterogeneity was evaluated with principal component analyses and outliers (>6 SD in any of the first 10 principal components) were removed. The criteria call rate, parameters of cluster plot irregularities according to Affymetrix’s recommendations, violation of Hardy-Weinberg equilibrium (*P*-value <10^-6^) in an exact test for autosomes, (*P*-value<10^-4^) for chromosome X with women only ^35^ and batch association (*P*-value<10^-7^) were considered during SNP QC. Subsequently, 7669 samples and 532,676 SNPs were imputed on the reference 1000 Genomes Phase 3 ^36^, applying SHAPEIT ^37^ v2r900 (prephasing) and IMPUTE2 ^37, 38^ v2.3.2 for genotype estimation. A specific genotype was assigned to a SNP if its corresponding genotype estimation featured a probability of >=0.8. In 2.5% of the cases, none of the genotypes exceeded that threshold and the respective SNP was labeled “missing” for that sample. SNPs whose “missing”-count over all samples exceeded upper quartile+1.5*IQR were removed resulting in a total of 2830 SNPs.

### matrixEQTL Analysis

Among samples with significant DMP data (from DMRs healthy *vs.* unhealthy), additional gene expression and SNP data was available for 48 samples. The R package matrixEQTL v2.3 ^39^ was employed on all three pairs of datasets in order to identify cis effects (within a range of +/-1kb) between methylation and expression (eQTMs), methylation and SNPs (meQTLs) and expression and SNPs (eQTLs). All three comparisons were performed on the complete data (*N*=48) and both subgroups with high LS (*N*=23) and low LS (*N*=25) separately. Since sex batch effects have been adjusted for in both expression and methylation data, small batch effects remained only for age and BMI. However, including both age and BMI as covariates into the matrixEQTL analysis did not change the overall result which is why the final matrixEQTL analysis was run without considering any covariances.

### Statistics

All statistical analysis were performed using *R software version 4.0.4* ^40^. After checking for normal distribution, Mann-Whitney U-Test or Welch’s t-test was applied to test for differences between 5^th^ and 95^th^ percentile as well as between lean and obese subgroups for the following phenotypes: BMI, age, HbA1c, waist-to-hip ratio (WHR), fasting plasma glucose and insuline, Low Density Lipoprotein (LDL)-, High Density Lipoprotein (HDL)-, apolipoprotein A1- and triglyceride serum levels. Welch’s t-test was used to compare methylation differences measured as normalized ß-values between low LS vs. high LS for each top DMP, respectively. Using first Shapiro-Wilk-test to proof normal distribution of the bisulfite sequencing data, independent Mann-Whitney U-Test was applied to compare methylation differences within the validation cohort. Methylation levels between BMI categories were compared applying 2-way ANOVA. Correlation analysis were performed using Spearman’s correlation. All respective analysis were adequately corrected for multiple testing.

## Results

### Self-reported lifestyle reflects obesity specific phenotypes

We correlated the Lifestyle Score (LS) with BMI and WHR in 4107 LIFE-Adult participants. The LS was related to the obesogenic environment (Supplemental Figure 4a-b) (all *P-*value<1x10^-3^) with significantly higher values in subjects with obesity (Supplemental Figure 4c). We furthermore demonstrated that all individual scores (diet, PA, smoking, alcohol and total LS) were mutually dependent (Supplemental Figure 4d), which was particularly marked in the extreme subgroups (5^th^ and 95^th^ percentile, Supplemental Figure 4e). Finally, our score showed simultaneous negative correlations (all *P-*value<1x10^-15^) to the protective lipid parameters HDL-C and apolipoprotein A1, which were higher in healthy living subjects (Supplemental Figure 5a-b).

When comparing healthy *vs.* unhealthy subgroups (5^th^ vs. 95^th^ percentile LS, healthy LS: *N*=216, unhealthy LS: *N*=207), nominal differences were observed regarding BMI distribution (Supplemental Figure 5, Table 1) (*N* lean/overweight/obese: healthy LS-73/95/48, unhealthy LS-67/93/46; *P*=0.051). Although HbA1c, fasting plasma glucose, fasting plasma insulin and LDL serum levels did not differ between the two groups either (all *P-*value >0.05, Table 1), strong differences were found for waist circumference (*P-*value <3x10^-6^), age, Apo A1 and triglyceride serum levels (all *P-*value <1x10^-9^) and especially HDL levels (*P-*value <1x10^-15^) and WHR (*P-* value <2.2x10^-16^). Subjects living an extreme healthy lifestyle were older (mean age difference = 6 years) with lower WHR (mean WHR difference = 0.08) and prominently lower lipid serum levels. However, as expected, all phenotypes differed significantly between subjects with and without obesity within healthy and unhealthy subgroups (all *P-*value <2x10^-2^) respectively, except for fasting plasma LDL. Regarding sex distribution, it is noticeable that females are overrepresented in the healthy subgroup, whereas males (Table 1) dominate the unhealthy subgroup.

### DNA methylation signatures are related to individuals’ lifestyle

By comparing genome-wide blood DNA methylation pattern in subjects with healthy vs. unhealthy lifestyle we identified 4682 significant DMRs annotated to 4426 genes with a min. smoothed FDR<5%, which included 220 DMRs with FDR<1x10^-4^ (Supplemental Table 4, Figure 1a). Amongst the significant DMRs the mean methylation level differences range from -6.9% to 5.5%. Given the rather subtle methylation changes for the majority of the DMRs, we introduced a mean methylation threshold of >2% to further narrow down the potential causal candidate DMRs. Among the 340 DMRs reaching this cut off, 164 DMRs were hypermethylated (mean methylation difference ±SD:2.6%±0.6%), whereas 176 DMRs were hypomethylated (mean methylation difference ±SD:-2.8%±0.8%) in healthy compared to unhealthy living individuals (Figure 1a, Supplemental Table 4). Taking into account a DMR can have more than one annotation, most DMRs (46%; counts relative to the number of DMRs) were located in CpG-islands, followed by 45% located in CpG-shores. In relation to gene regions, most DMRs are located in introns (59%), followed by exons (39%) (Figure 1b).

**Figure 1.**
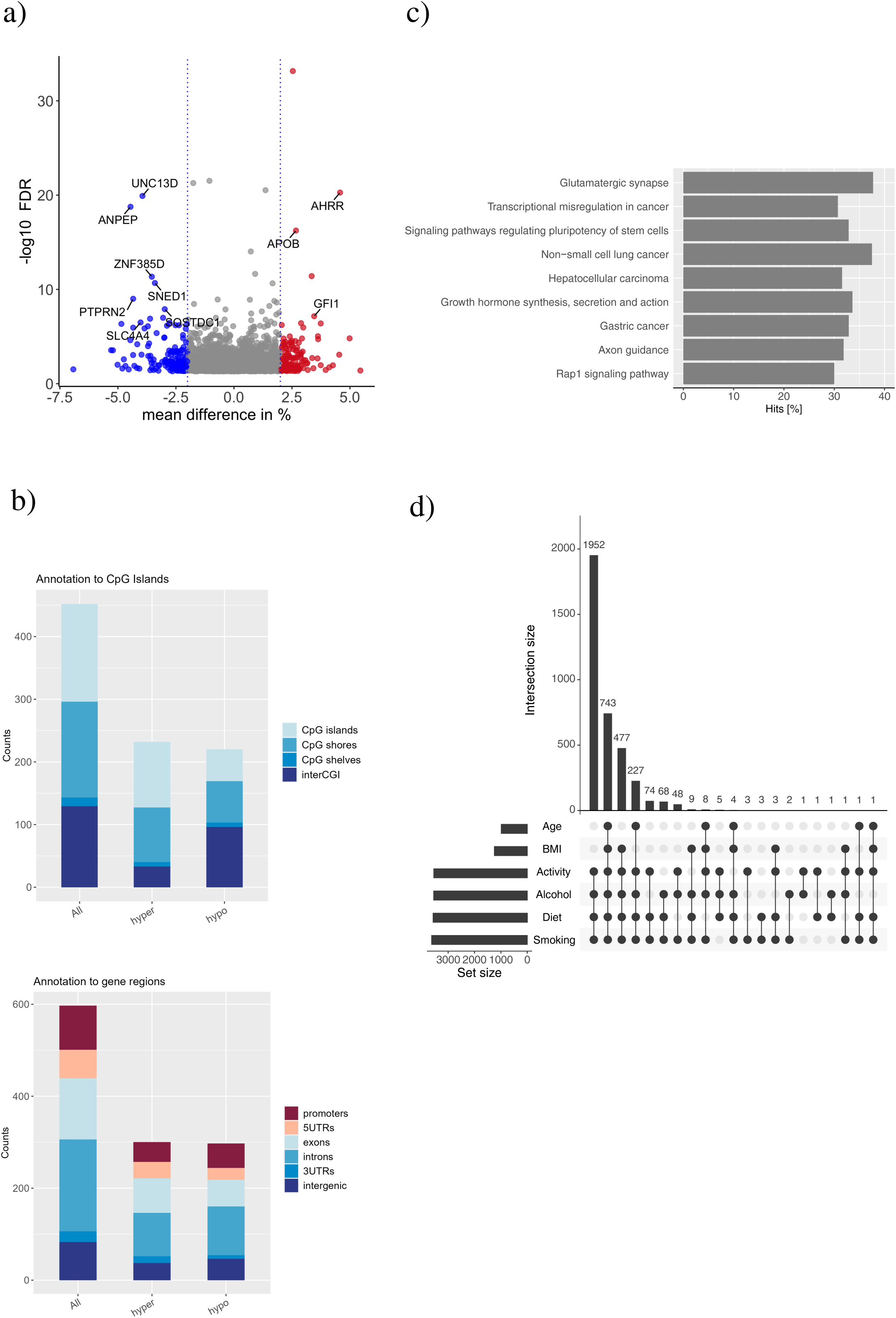
Lifestyle-specific Differentially Methylated Regions (DMRs) in the discovery cohort: a) Volcano plot representing the significant DMRs (min. smoothed FDR<5%) based on the healthy *vs.* unhealthy lifestyle comparison. Positive mean methylation differences ≥|2|% represent hypermethylated DMRs (red dots) and negative methylation difference ≥|2|% hypomethylation DMRs (blue dots) in the healthy subgroup. b) The location of the DMRs in relation to CpG islands (top) and the location of the DMRs in relation to gene regions (bottom). Both plots are presented as number of counts including multiple annotations. Hyper: hypermethylation, hypo: hypomethylation. c) KEGG pathway enrichment analysis presented as percentage of annotated genes relative to all genes involved in the respective pathway (hits in %) for all enriched pathways with an FDR<5%. d) Intersection plot illustrating the frequency of significant DMRs driven by any of the included lifestyle aspect (diet, physical activity, smoking and alcohol) and the potential confounders age and BMI. The majority of the DMRs are driven by an interaction between all four lifestyle aspects.

The top 15 hypo- and hyper-methylated significant DMRs according to their mean methylation difference are presented in Table 2 with a DMR annotated to the *Glutamine-Fructose-6-Phosphate Transaminase 2* (*GFPT2*) gene locus showing the strongest hypomethylation (mean methylation difference DMR=-6.9%). A DMR annotated to the *Glutamate Rich 1* (*ERICH1*) gene showed the strongest hypermethylation (mean methylation difference DMR=5.4%). Finally, using KEGG pathway analyses we identified the glutamatergic synapse as the most enriched pathway (adj. *P*-value<0.01) followed by axon guidance, another brain related pathway (adj. *P*-value <0.05). Most of the nine enriched pathways (Supplemental Table 5 and Figure 1c) are related to various cancer types.

As demonstrated by the intersection plot (Figure 1d, Supplemental Table 6) the majority of the DMRs (*N*=1952) are driven by all 4 lifestyle subscores together (diet, PA, smoking and alcohol), followed by a combination of them together with BMI and age *(N*=743). Obviously, BMI and age alone do not explain any of the identified DMRs. Although, this did not indicate a prominent role of smoking (Figure 1d), given by the nature of the lifestyle score a comparison between participants with very healthy and very unhealthy lifestyle mirrors differences between non-smokers and smokers. We therefore further adjusted the complete analyses for smoking as covariate, which resulted in 629 identified DMRs with a min. smoothed FDR<5%. Amongst them, the most significant DMR is located within a CpG Island of the *Ring Finger Protein 39* (*RNF39*) locus (Supplemental Table 7).

### Lifestyle-derived DMPs correlate with metabolic traits related to obesity

DMP-specific analysis comparing subjects with healthy and unhealthy lifestyle identified 145 significant DMPs (adj. *P*-value<5%) (Figure 2a, Supplemental Table 8). A total of 26 DMPs passed a mean methylation change |(logFC)| ≥5%, 14 of which were hypermethylated logFC≥5% (0.07±0.04) while 12 showed hypomethylation logFC≤-5% (-0.06±0.01). Of these, 19 DMPs were clearly assigned to a specific gene. However, when considering significance levels (adj. *P-value<5%*) as well as mean methylation change ≥5%, the strongest effects were observed for *AHRR* (Aryl-Hydrocarbon Receptor Repressor), *F2RL3* (*F2R Like Thrombin or Trypsin Receptor 3*), *RARA* (*Retinoic Acid Receptor Alpha*), and *PRSS23* (Serine Protease 23) (Figure 2b, all *P*-value<1x10^-11^, Supplemental Table 8), all of them being hypomethylated under unhealthy conditions. Correlation analysis between the identified DMPs and obesity related phenotypes (HbA1c, LDL, HDL, blood glucose, insulin, apolipoprotein A1, triglycerides, BMI, waist circumference, WHR and age) as well as the LS and its subscores can be found in Supplemental Table 9. Two of the 20 included DMPs showed significant correlation with WHR even after Bonferroni correction, although no SNPs within the genomic region have been shown to be associated for BMI-adjusted WHR in previous GWAS ^41^ (GWAS catalog accessed June 2021). Amongst the identified DMPs the strongest correlation to WHR was found for *Vasohibin 1* (*VASH1*) (*P*-value=4.2x10^-5^, r=-0.4), which is in line with an association to HbA1c (%) (*P*-value=0.03, r=-0.21), triglycerides (mmol/l) (*P*-value=0.046, r=-0.2) and waist circumference (cm) (*P*-value=0.04, r=-0.2), although only the association with WHR sustained after correction for multiple testing. Finally, two DMPs of *F2RL3*, one of our selected top hits, showed marginal correlations to LDL serum levels (all *P*-value<0.03; r=-0.22) and triglycerides (mmol/l) (all *P*-value<0.07, r=-0.27) and one DMP additionally to HbA1c (%) (*P*-value<0.01, r=-0.3). The selected top DMPs for *F2RL3* showed additional correlations to HDL (mmol/l) (*P*=0.03, r=0.23) and apolipoprotein A1 (g/l) (*P*=0.03, r=0.23), albeit not withstanding corrections for multiple testing.

**Figure 2.**
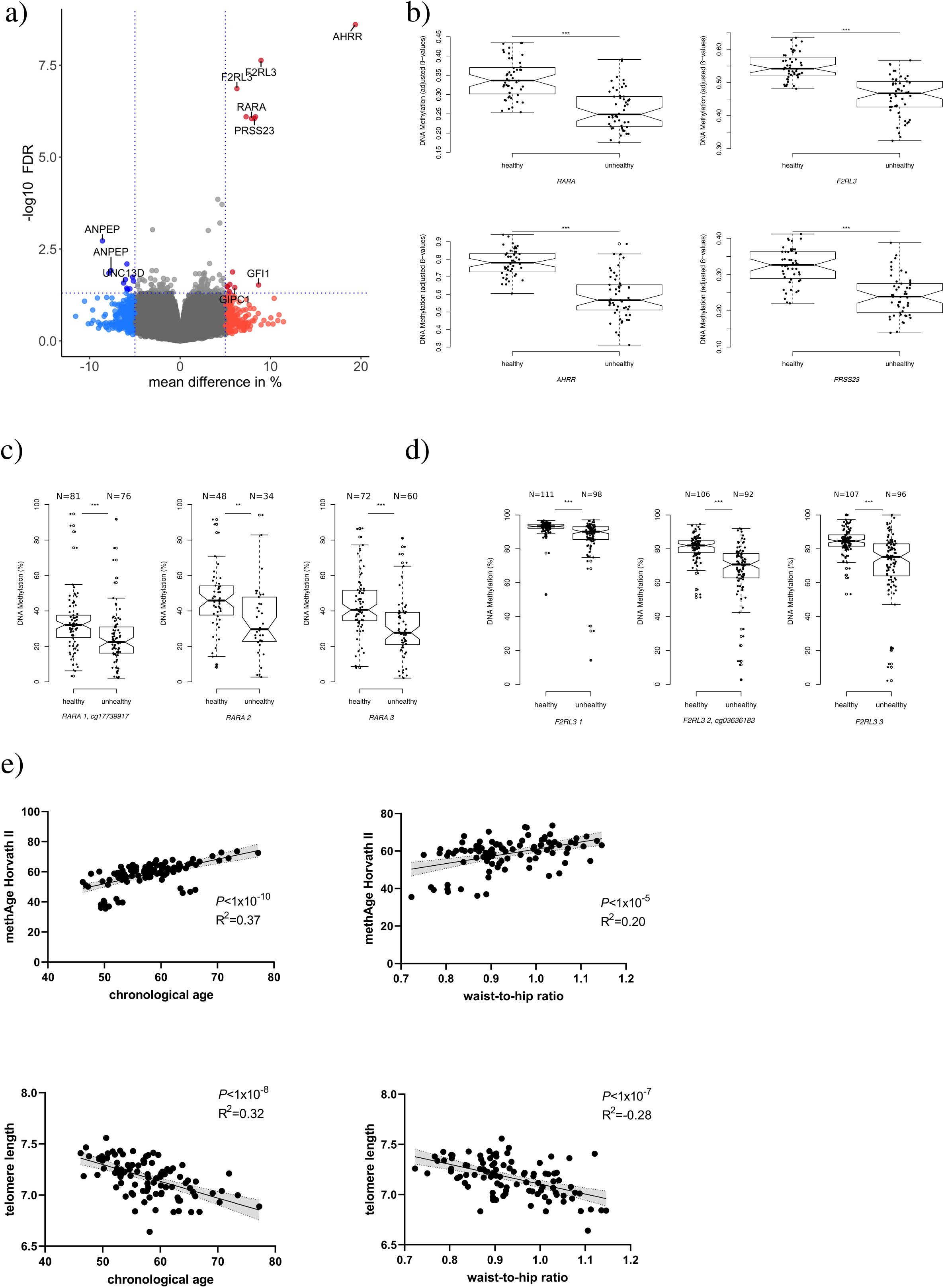
Differentially Methylated Positions (DMPs) comparing healthy vs. unhealthy lifestyle in the discovery (a-b) and the validation cohort (c-d): a) Volcano plot representing the significant DMPs (min. smoothed FDR<5%) based on the healthy *vs.* unhealthy lifestyle comparison. Positive mean methylation differences ≥|5|% represent hypermethylated DMPs (red dots) and negative methylation difference ≥|5|% hypomethylation DMPs (blue dots) in the healthy subgroup. b) Box plots representing mean methylation±SD for the top four identified genes: *RARA* (cg17739917); *F2RL3* (cg03636183); *AHRR* (cg05575921); *PRSS23* (cg14391737) comparing healthy (low Lifestyle Score (LS)) vs unhealthy (high LS) living subjects, 95% confidence interval is represented by notches. *P*-values indicate statistically significant differences detected using Welch’s t-test. c-d) Box plots are given as mean methylation±SD as well as the 95%-confidence interval is represented by notches for the two validated DMPs (c) *RARA* and d) *F2RL3)* and their surrounding CpGs. *P*-values indicate statistical significance between healthy (low LS) and unhealthy (high LS) subjects detected using ANOVA. *P*-values are indicated as: *<0.05, **<0.01 and ***<0.001 e) Linear regression analysis between methyl age (methAge) for the Horvarth II, telomere length, chronological age and Waist-to-Hip Ratio (WHR) measurements presented as scatter plot. The light grey area represents the 95% confidence interval and R^2^ representing the coefficient of determination.

In summary, 13 of the 20 selected DMPs showed marginal correlations with triglycerides (all *P*-value<0.046), 8 with HDL (all *P*-value<0.047) and 7 with LDL (all *P*-value<0.033) serum levels (Supplemental Table 9).

### Bisulfite sequencing within RARA and F2RL3 loci supports findings from the discovery stage

Based on these findings and further supported by previously reported data ^42–44^, we selected two DMPs (*F2RL3:cg03636183* and *RARA:cg17739917*) for validation (*N*=213) using bisulfite sequencing and demonstrate directionally consistent differences between subjects with healthy vs. unhealthy lifestyle (Figure 2c-d; both *P*-value<0.001, mean meth. diff.: *RARA* CpG=8.37%, *F2RL3* CpG2=13.87%). Furthermore, the surrounding CpG position confirmed this effect with all *P*-value<0.01 and a mean methylation difference of >5.59% (mean meth. diff.: *RARA* CpG2=11.99%, *RARA* CpG3=12.58%, *F2RL3* CpG1=5.59%, *F2RL2* CpG3=13.2%) (Figure 2c-d). The here used validation cohort did not differ significantly from the discovery cohort regarding age, BMI and sex distribution (Table 1). Spearmańs correlation analysis showed a significant correlation between methylation levels observed in the genome-wide methylation analysis and bisulfite sequencing (*RARA*: spearmańs r=0.45, *F2RL3*: spearmańs r=0.53, both *P*<7.9x10^-5^) (Supplemental Figure 5c-d).

### Obesity specific methylation marks

Driven by the comparable distribution of subjects with and without obesity between very healthy and unhealthy lifestyle groups, we aimed to identify lifestyle independent obesity-related methylation marks. Therefore, blood methylation patterns of subjects with obesity (*N*=25) were compared with subjects without obesity (*N*=25) within each lifestyle group separately. Interestingly, whereas about 1572 DMRs annotated to 1599 different genes were identified in healthy subjects, only 85 DMRs annotated to 101 genes were detected in subjects living an unhealthy lifestyle (Supplemental Table 10 and 11) with a min. smoothed FDR<5%. This further included 10 identical annotations amongst the *PAX6* and *HOXA9-10* gene clusters, already known candidates regarding obesity and related co-morbidities. However, on CpG levels no DMPs sustained after correction for multiple testing (data not shown). Nevertheless, KEGG pathway analysis for the healthy subgroup indicated eight enriched pathways, amongst them GABAergic synapse, dilated cardiomyopathy and calcium signaling (Supplemental Figure 6, Supplemental Table 12), whereas for the unhealthy subgroup only antigen processing and presentation was enriched (not shown).

### Methylation Age

We observed the strongest association (*P*-value<1x10^-10^, R^2^=0.37, Figure 2e) between methylation age and subjects’ chronological age within the discovery cohort for the Horvath’s clock II, which was compared to Horvath’s I, additionally trained on 850K EPIC arrays (Supplemental Figure 7a-c). Only marginal (*P*-value=0.01) differences in DNAmAge acceleration were observed when comparing individuals with healthy vs. unhealthy lifestyle, which was similar to comparing never smokers with previous or current smokers (Supplemental Figure 7d and f). No difference was observed between subjects with and without obesity (Supplemental Figure 7e). We furthermore observed a strong negative association between the telomere lengths clock and chronological age (*P*-value<1x10^-8^, R^2^=-0.32, Figure 2e). Interestingly, both clocks showed an additional linear association to WHR within our discovery cohort (all *P*-value<1x10^-4^, Figure 2e).

### Underlying genetic predispositions and effects on mRNA level in blood

Driven by the small overlapping sample size (*N*=48) and only marginal genetic variation in close proximity (+/1 kb) to the identified target DMRs (healthy vs. unhealthy lifestyle) we could not identify any meQTLs or eQTLs. However, we found associations between methylation levels of 8 DMPs with target mRNA expression levels (Supplemental Table 13) in the combined discovery group (all individuals with healthy and unhealthy lifestyle). Amongst them, only two eQTMs annotated to the *ANPEP* (*Alanyl Aminopeptidase*) locus sustained after correction for multiple testing (matrix FDR=0.03; Supplemental Table 13; Supplemental Figure 8). Four eQTMs were detected in subjects with healthy and 8 with unhealthy lifestyle, amongst them also one of our candidate DMP of *F2RL3* in healthy subjects.

## Discussion

Epigenetic markers are known to reflect environmental conditions and thereby are not only affected by genetic predisposition, but most strongly by our daily lifestyle. Although this is widely acknowledged by the scientific community, the majority of epigenetic studies in regard to obesity, most of them conducted cross-sectionally, are still lacking the inclusion of relevant lifestyle drivers^45^. Therefore, to the best of our knowledge, this is one of the few studies investigating potential effects of lifestyle on the respective blood DNA methylation signatures ^46^. Here, we calculated LS scores based on each individuals’ diet, physical activity, smoking, and alcohol consumption within the LIFE-Adult study from Germany. Genome-wide DNA methylation analysis in blood samples of 100 subjects representing healthy and unhealthy lifestyle extremes demonstrated that daily lifestyle is most likely superior to the obesity state itself in associations with blood DNA methylation pattern, as supported by association studies between neonatal blood methylation and the risk to develop obesity later in life ^47^. The study showed that the distribution of obesity categories in extreme lifestyle groups was comparable, and that potentially obesity-associated methylation marks were more frequent in subjects with healthy lifestyle. Furthermore, methylation age and estimated telomere length showed strong correlations to chronological age and WHR, with observed smaller DNAmAge acceleration distances in healthy subjects. Finally, two DMPs for *ANPEP* also showed the strongest eQTM in blood.

With this study, we took several lifestyle aspects into account to explore relations between long-term lifestyle habits and differences in human blood methylation patterns. Our findings imply that dietary habits, physical activity, smoking habits and alcohol consumption are influencing epigenetic patterns together, whereas only neglectable effects are attributed to age and BMI alone. This suggests that rather than simply representing the consequence of obesity, differences in blood derived methylation marks may be primarily driven by long-term lifestyle habits. This is further supported by observed smaller DNAmAge acceleration in healthy compared to unhealthy lifestyle group, whereas no significant difference could be observed between subjects with and without obesity.

We identified several candidate genes differentially methylated according to the LS and successfully validated *RARA* and *F2RL3*, already known from previous studies to be influenced by lifestyle aspects and acknowledged for their role in metabolic diseases ^42, 44, 48^. Both genes were hypermethylated within extremely healthy compared to unhealthy living individuals, which is in line with previously published data ^42, 49, 50^. In particular, hypomethylation of the DMP within the *F2RL3* locus appears to increase the risk for cardiovascular as well as overall mortality ^42, 49^. Translated to our results this might indicate an increasing mortality risk of an unhealthy lifestyle accompanied with associated diseases such as obesity, type 2 diabetes, cardiovascular diseases or cancer. Previous studies further showed a hypomethylating effect of smoking on the here identified *F2RL3* DMP ^48^. Moreover, very recently a strong association to coffee consumption in a large-scale EWAS was reported ^51^. Consistent with published data on smoking, subjects with unhealthy lifestyle in our study showed a mean methylation of 67% compared to 81% in the healthy lifestyle group, with the majority of subjects within the unhealthy group being actual smokers (validation cohort). In line with this we further observed a marginal (*P*-value=0.04) positive correlation between this methylation of this DMP and *F2RL3* mRNA levels in the healthy lifestyle subgroup. We found significant methylation differences between the healthy and unhealthy lifestyles for *RARA*, known for its role in adipogenesis ^52^. It is noteworthy though, that based on the findings of the present study, an increase in *RARA* methylation pattern might be related to higher HDL-C and lower triglyceride serum levels, indicating a link between RARA and lipid metabolism.

Although the observed differences in DNA methylation in *RARA* could also be driven by smoking as previously described ^43^ and supported by the here found strong correlation with smoking, there is still a prominent influence of other environmental conditions like diet and physical activity as shown by our present data. Nevertheless, it needs to be acknowledged that in line with our study, the majority of methylation studies on smoking, although lacking any information on diet or activity, identified a similar set of top candidates especially including *F2RL3*, *RARA* and *AHRR* in human blood cells ^42, 44, 49, 50^. It is also worth to mention, that smoking effects on *F2RL3* methylation were previously also observed in adipose tissue ^44^. Consequently, narrowing down our list of potential lifestyle discriminating candidates regions by including an additional adjustment for smoking resulted in identification of a top DMR on chromosome 6 annotated to the *Ring Finger Protein 39* (*RNF39*). This DMR is overlapping a larger region very recently described to successfully discriminate responder from non-responder to an lifestyle intervention based on either Mediterranean/low-carbohydrate or low-fat diet with or without physical activity ^7^.

There are a few key limitations to our study. First, we used a scoring system based on self-reported questionnaires, which might lead to euphemistic information including over- or underestimation of the real status ^53^. However our study design is supported and strengthened by findings, which are in line with previously reported data e.g. on lifestyle factors such as smoking ^42, 44^. Finally, the observational and cross-sectional nature of the study does not allow testing the direction of causality at least between methylation and metabolic phenotypes and limits our ability to rule out confounding (e.g. sex), even though it seems unlikely that methylation marks affect lifestyle habits. Although the identification of reliable and reproducible epigenetic marks for obesity in human blood remains challenging, our study clearly indicates the importance of considering as many lifestyle aspects as possible when analysing epigenetic data with regard to complex diseases like obesity. We successfully demonstrate that the majority of CpG methylation marks are much stronger influenced by our daily lifestyle than the obesity state itself.

## Data Availability

All data generated or analyzed during this study are included in this published article and its supplementary information files. Raw data will be made available via the Leipzig Health Atlas after acceptance of the publication.

## Acknowledgements

We thank all study participants of the LIFE Adult study whose personal dedication and commitment have made this project possible. We would like to acknowledge excellent technical assistance by Beate Gutsmann and Ines Müller.

## Author contributions

MK, PK, PWF and MSc initiated, conceived and designed the study. IK and MK performed lifestyle analysis, selected samples and wrote the first manuscript draft. IK performed DNA quali-and quantification and performed most of the laboratory work. AH, IK, TH, LM and MK performed statistical and bioinformatic analysis. MSc, RB, JT and ML conducted the LIFE population based study, collected the phenotypes and generated genome and transcriptome data. LM, KR, AT, YB, MS, MB, MSc and PWF supported the critical data interpretation and reviewed the manuscript. AH, PK and MK wrote the final manuscript version.

## Consent for publication

Not applicable

## Supplemental Figures

**Supplemental Figure 1.**
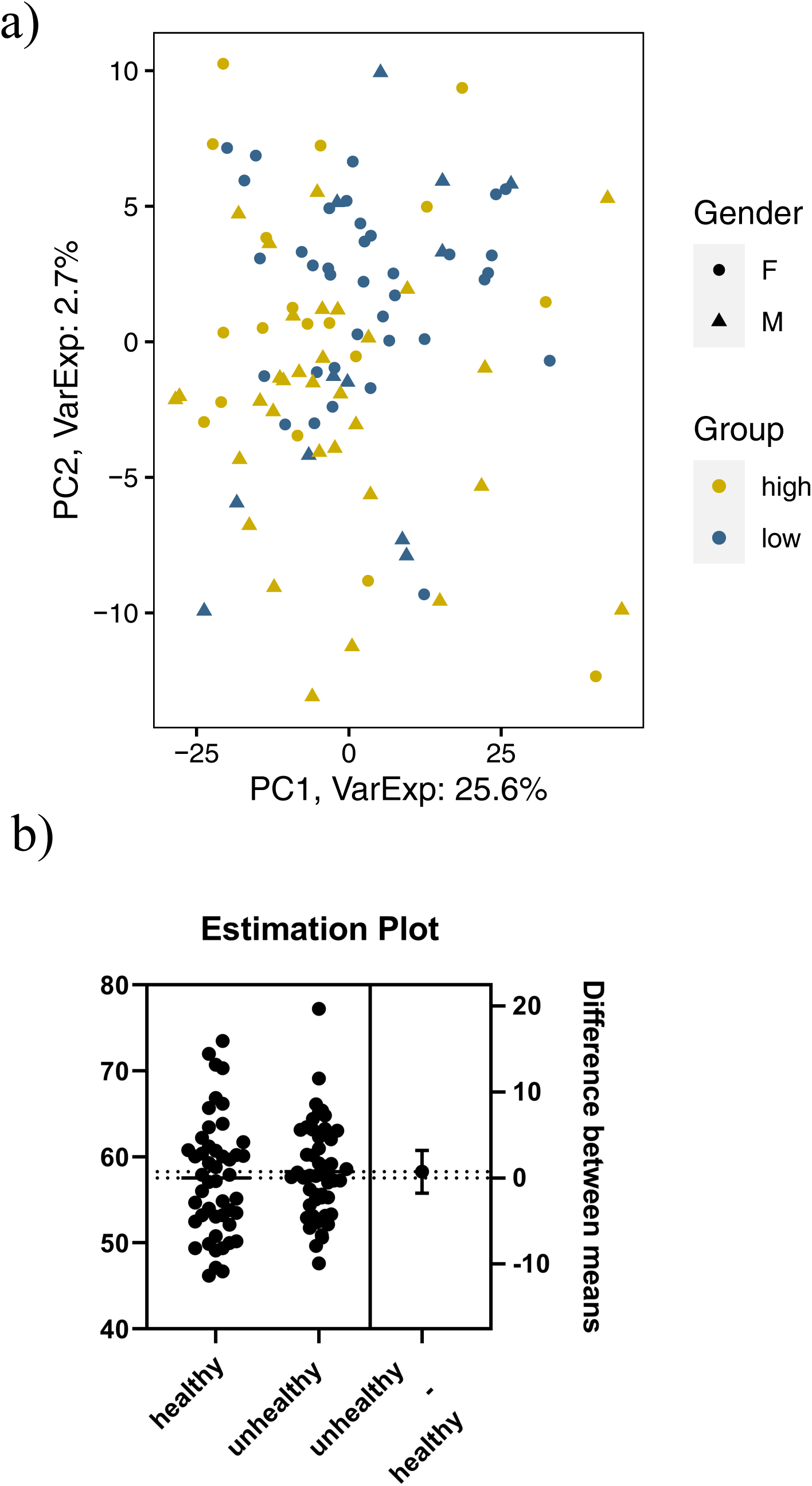
sex and age specific quality control: a) Principal Component Analysis (PCA) of the Illumina 850K methylation arrays. After sex normalization, no batch effect for sex is visible anymore, but the separation for the healthy (high) and unhealthy (low) living subjects is evident. b) Estimation plot using an unpaired t-test for age differences between low vs. high LS (healthy vs. unhealthy) subgroups.

**Supplemental Figure 2.**
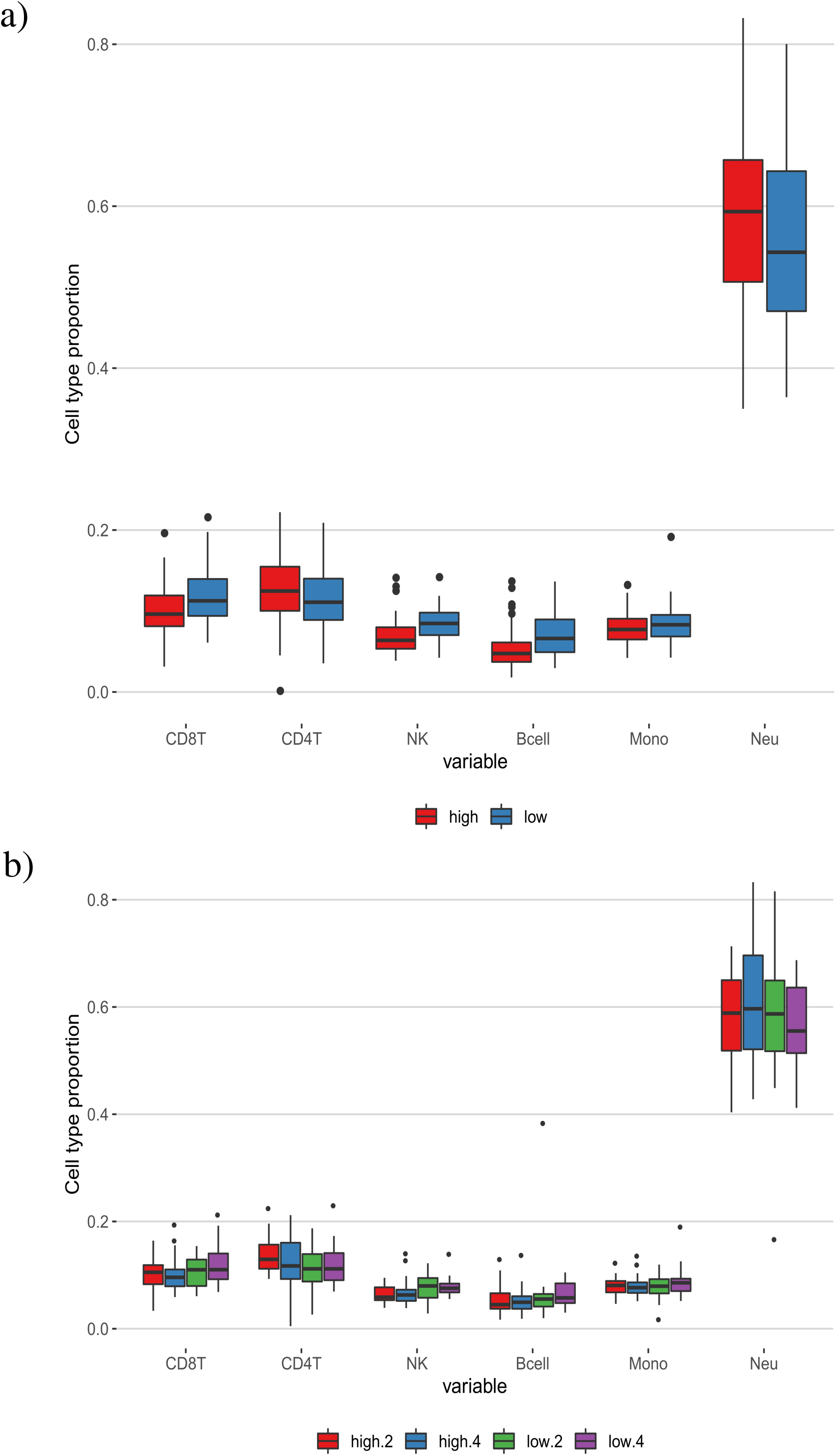
Cell type distribution: Box plots representing the cell type proportions for a) low and high Lifestyle Score (LS) subgroups, and b) for low and high LS subgroups in individuals without (2) and with obesity (4) based on Illumina 850K methylation data. CD8 T- lymphocates (CD8T), CD4 T-lymphocytes (CD4T), natural killer cells (NK), B-lymphocytes (Bcell), monocytes (Mono) and neutrophils (Neu).

**Supplemental Figure 3.**
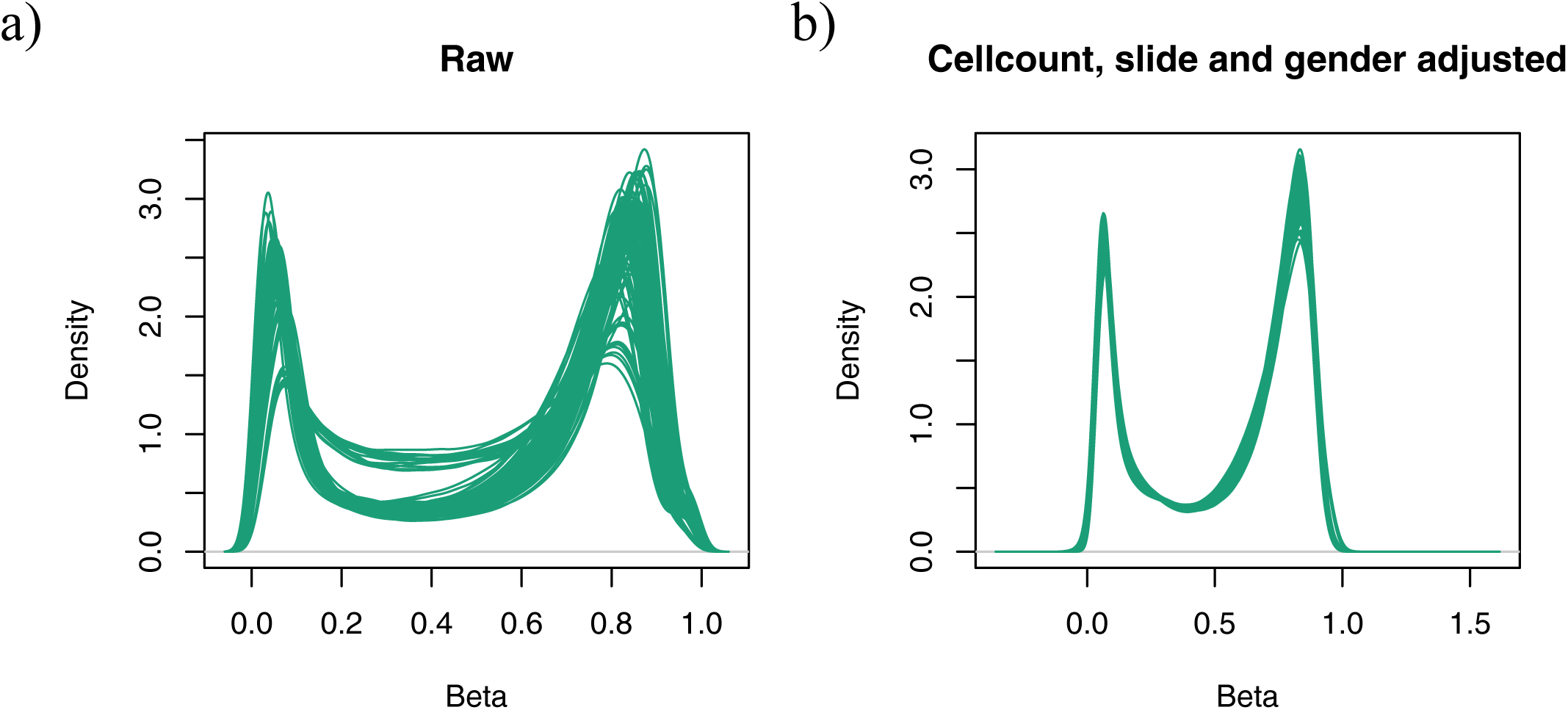
*ß*-values densities of the Illumina 850K methylation data: The densities of *ß*-values are shown for a) the raw data and b) after adjustment for cell counts, array slides and gender. Since the arrays were run on two batches, differences are visible in the raw data and had to be corrected for.

**Supplemental Figure 4.**
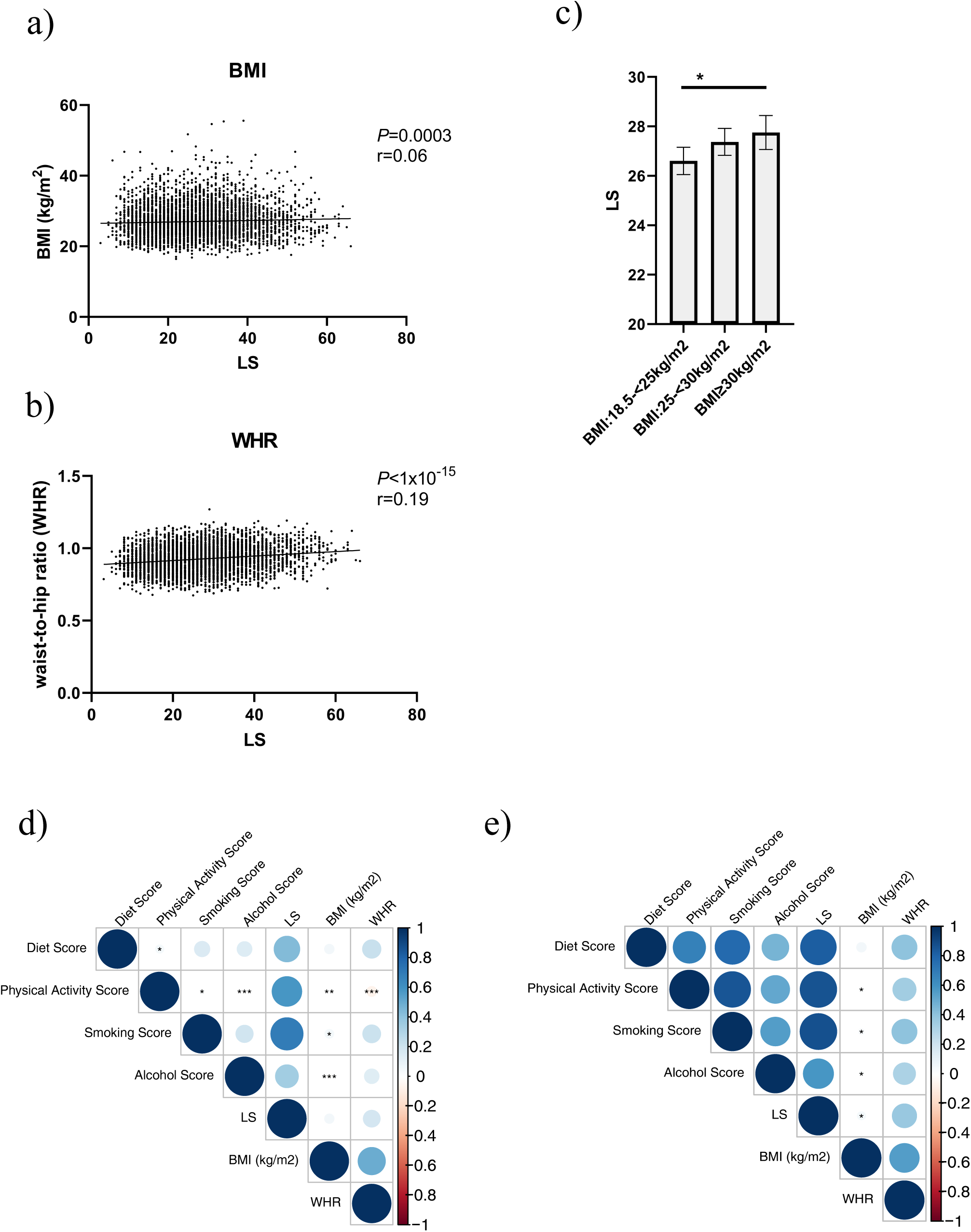
**Lifestyle Score (LS) associations in the LIFE cohort**: Scatter dot plots representing spearman correlations between the LS and a) BMI in kg/m^2^, b) Waist-to-hip ratio (WHR) for the total cohort (N=4107). c) Bar plot representing the LS values as mean±SEM for defined BMI categories. Intergroup differences were assessed using student’s t-test; *: *P*<0.05. d-e) Correlations between lifestyle scores (including sub scores) are shown. LS and anthropometric measurements are presented as correlation matrix. Color and size of the dots represent Spearmańs correlation coefficient r; p-values are indicated with ***: *P*<0.001, **: *P*<0.01, *:*P*<0.05. Figure d) total cohort (N=4107), e) significant correlations within the sub scores in the validation cohort consisting of the extreme lifestyle edges (N=213).

**Supplemental Figure 5.**
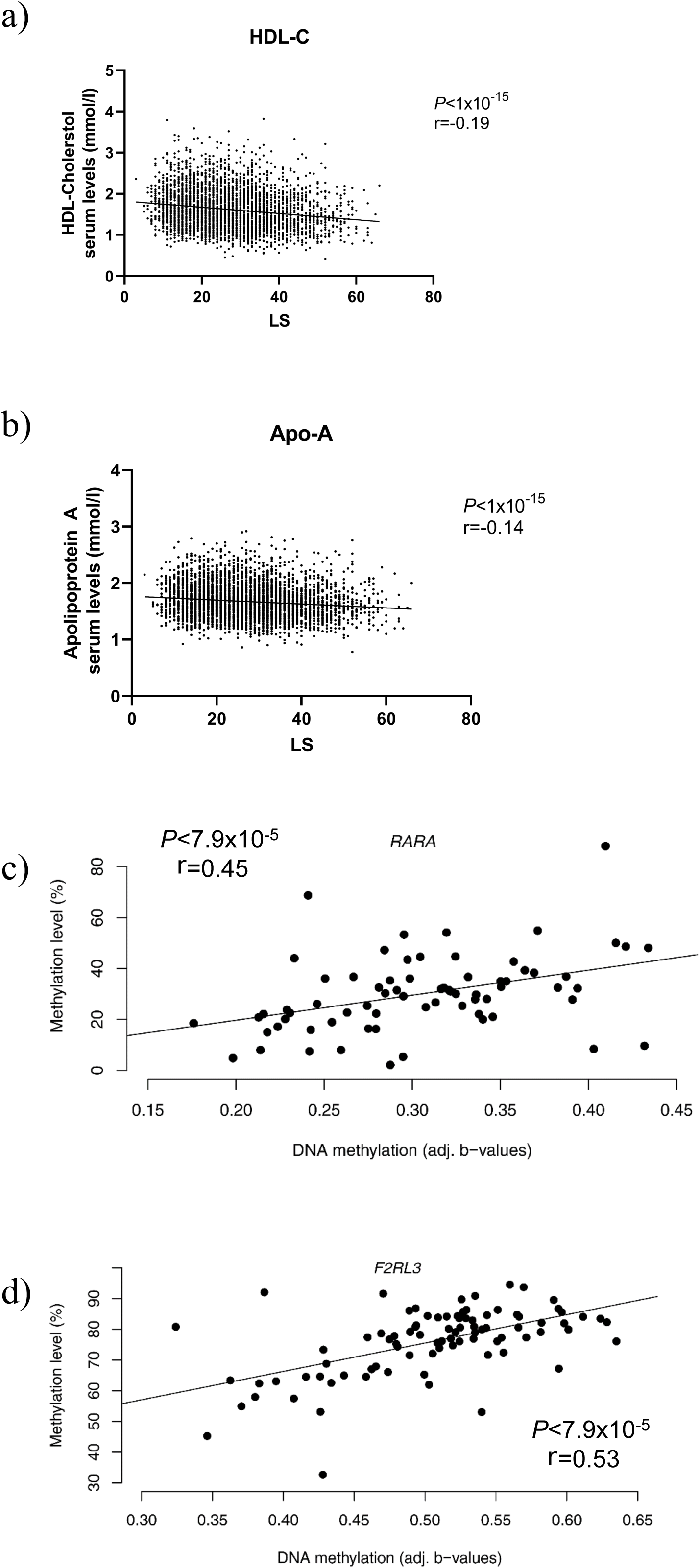
Lifestyle Score (LS) correlations in the total LIFE cohort (*N*=4107): Scatter plots representing Spearman’s correlations between the LS and a) High Density Lipoprotein (HDL)-Cholesterol serum levels (mmol/l) and b) Apolipoprotein A serum levels (mmol/l). Spearmańs correlation analysis between methylation levels observed in the Illumina EPIC arrays and the pyrosequencing sequencing validation are shown in c) for the *RARA* and d) for the *F2RL3* identified CpG position.

**Supplemental Figure 6.**
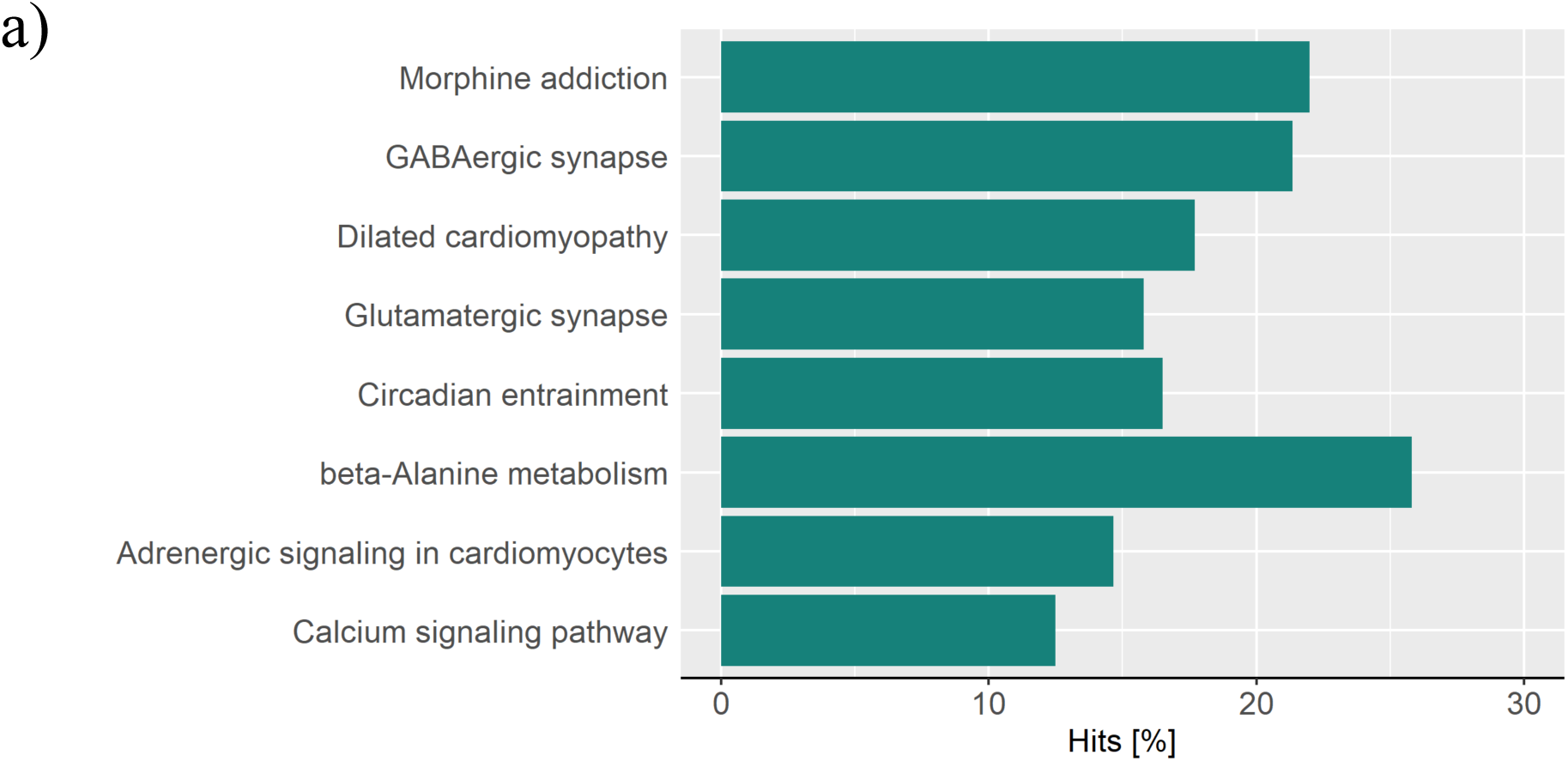
KEGG pathway enrichment analysis of the healthy subgroup: BMI related (non-obese vs. obese) KEGG pathway enrichment in the low **Lifestyle Score (**LS) subgroup is presented as hits in % for all pathways with an FDR<0.05.

**Supplemental Figure 7.**
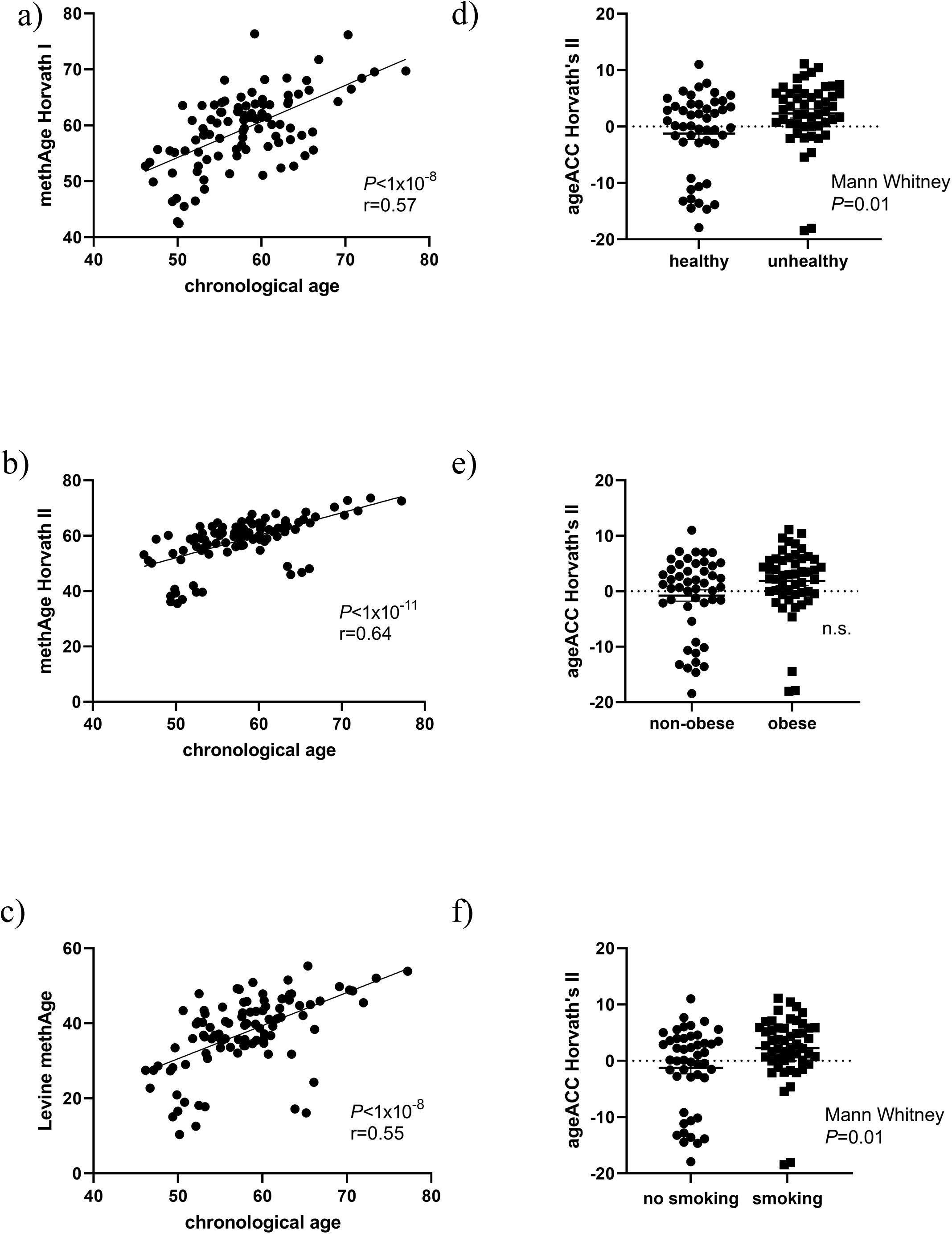
Methyl age (MethAge) correlations in the discovery cohort: Spearman correlation between MethAge for a) Horvath I, b) Horvath II and c) Levine’s clock and individuals chronological age in the discovery cohort (*N*=100) presented as scatter dot plot. d-f) Plots presenting differences in DNA methylation age (DNAmAge) acceleration as mean±SEM between d) healthy and unhealthy living subjects, e) subjects without and with obesity and f) never- vs. current/previous smokers.

**Supplemental Figure 8.**
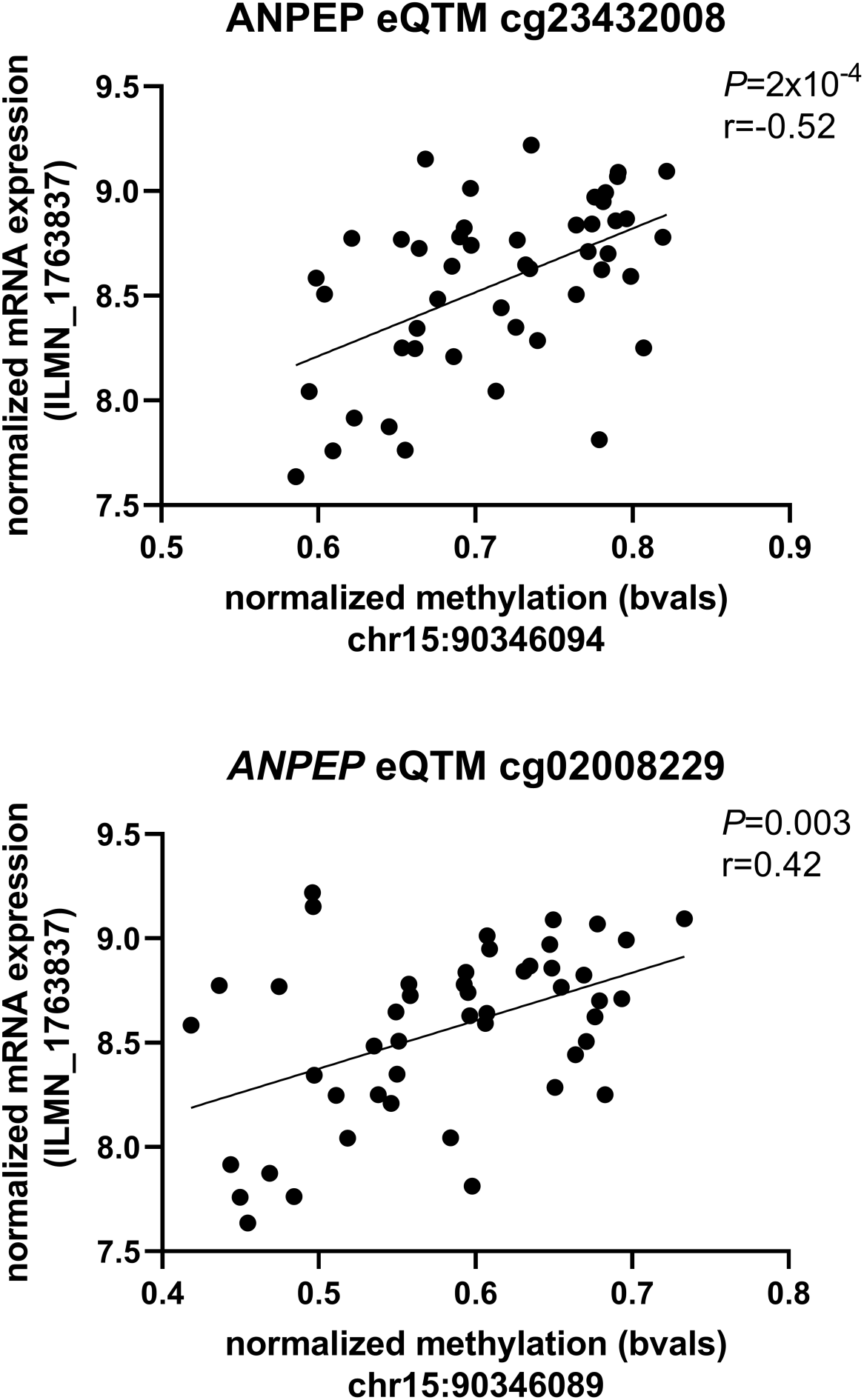
Spearman’s correlation between normalized methylation for *ANPEP DMPs* (cg23422008 and cg02008229) and corresponding normalized mRNA expression. (ILMN_1763837) presented as scatter plot.

## Supplemental Tables

**Supplemental Table 1 -Detailed Scoring System**: Diet Score was assessed using FFQ-information, favorable food intake got decreased scoring points with increased frequency, unfavorable food intake reached higher scores with regular consumption (high diet scores means unhealthy nutrition intake). Physical activity was rated via activity categories and MET-min/week divided into quartiles, counted by the SF-IPAQ (less active means higher scores). Smoking assessment is based on actual smoking status and information about pack years (higher nicotine consumption means higher scores). Alcohol consumption was considered harmful by the cut-offs based on DGE recommendations (exceeded consumption means higher scores).

**Supplemental Table 2 -Study Characteristics of the subgroups:** Phenotypic data is described for discovery and validation cohort subjects within the healthy (≤5^th.^ percentile) and unhealthy (≥95^th^ percentile) living extreme subgroups as mean±SD.

**Supplemental Table 3 -Self-designed primer sequences:** Self-designed primer sequences were used to validate candidate CpGs (*F2RL34* and *RARA*) with pyrosequencing technique, based on the PyroMark Q24 platform.

**Supplemental Table 4 – All lifestyle-specific Differentially Methylated Regions (DMRs):** Overview of all significant (min. smothed FDR<5%) DMRs of the healthy vs. unhealthy lifestyle analysis.

**Supplemental Table 5 – Lifestyle-specific KEGG pathway enrichment analysis:** KEGG pathway overrepresentation analysis for Differentially Methylated Regions (DMRs) from the healthy vs. unhealthy Lifestyle-Score analysis. All significant DMRs were included. Enrichment *P*-values were adjusted using Benjamini-Hochberg correction and FDR<5% were considered as statistically significant.

**Supplemental Table 6 -Differentially Methylated Regions (DMRs) of the lifestyle subscores:** Overview of all significant DMRs of the healthy vs. unhealthy lifestyle analysis including individual adjustments for confounder (BMI and age) and all subscores (diet, physical activity (PA), smoking and alcohol.

**Supplemental Table 7 – Lifestyle-specific Differentially Methylated Regions (DMRs) after smoking adjustment:** Overview of all significant DMRs from the healthy vs. unhealthy lifestyle analysis with a min. smoothed FDR<5% after additional adjustment for smoking score are shown.

**Supplemental Table 8 -All lifestyle-specific Differentially Methylated Positions (DMPs):** Overview of all statistically significant (adj. *P*-value<0.05) DMPs from the healthy vs. unhealthy lifestyle study are given. Our selected candidates (*F2RL34* and *RARA*) for the bisulfite validation are highlighted.

**Supplemental Table 9 -Lifestyle-specific Differentially Methylated Positions (DMPs) correlation analysis:** Correlation analyses between obesity-related phenotypes and the top 10 hyper- and hypomethylated DMPs between healthy vs. unhealthy lifestyle within the discovery cohort are given. *P*-values maintaining after correction for multiple testing [adj. *P*-values<1.5x10^-4^; 0.05/(number included DMPs*number obesity-related phenotypes)], are indicated in bold.

**Supplemental Table 10 - Differentially Methylated Regions (DMRs) of subjects with healthy lifestyle:** Overview of all significant DMRs within subjects with healthy lifestyle comparing non-obese vs. obese phenotypes.

**Supplemental Table 11 - Differentially Methylated Regions (DMRs) of subjects with unhealthy lifestyle:** Overview of all significant DMRs within subjects with unhealthy lifestyle comparing non-obese vs. obese phenotypes.

**Supplemental Table 12 - KEGG pathway enrichment analysis of subjects with healthy lifestyle:** KEGG pathway overrepresentation analysis for Differentially Methylated Regions (DMRs) from non-obese vs. obese phenotypes in subjects with extreme healthy lifestyle. All DMRs with FDR<5% were included. Enrichment *P*-values were adjusted using Benjamini-Hochberg correction and only hits with an FDR<0.05 were considered statistically significant.

**Supplemental Table 13 -Expression Quantitative Trait Methylation Analysis (cis-eQTM):** cis-eQTM analysis for the combined discovery cohort and the healthy- and unhealthy lifestyle groups is given. All DMPs with a *P*-value<0.05 are listed including the corresponding T-test statistic and beta values as effect size estimates according to matrixEQTL, gene locus, strand and UCSC annotation. The associated expression probe is listed with its gene locus and annotation according to Illumina. The DMP *P*-values were adjusted using Benjamini-Hochberg correction and only hits with an FDR<5% were considered statistically significant and are highlighted in bold

## Supplemental Material

### Lifestyle sub-scores

Briefly, the diet score was based on the German FFQ where all participants were asked about their frequency in consumption of 34 different food groups and 13 beverages during the last year^1^. Among them we considered six food groups as favorable (e.g. fish, vegetables, and whole grain products) and eight as unfavorable (e.g. pan-fried potato products, meat and cold cuts, fast-food, and instant products), after testing their correlation to BMI. All Participants could choose from several categories: eating each food group a) several times per day, b) each day or almost each day, c) several times per week, d) once per week, e) two or three times per month, f) once or less a month, g) or rather never^2^ (Supplemental Table 1). For the diet score, frequently consumed unfavorable foods counted high, whereas frequently consumed favorable foods counted low (Supplemental Table 1). In line with this, a high diet score correspond to an unfavorable eating pattern as previously described^3^.

For the physical activity score we used the SF-IPAQ^4^ and included the absolute activity measured as Metabolic Equivalent of Task-minutes per week (MET-min/week) and the categorical physical activity level as a) low, b) moderate, and c) intensive, as results of the questionnaire evaluation algorithm^1,5^. We scored the activity levels from intense to moderate to low (0-5-10) and used quartiles from the entire cohort distribution of MET-minutes/week for the absolute activity scoring as shown in Supplemental Table 1. In total, the physical activity score was calculated as sum of absolute and categorical activity levels.

For smoking behavior, we used the available data from the LIFE-Adult cohort regarding the current smoking status (non-smoker, previous smoker and current smoker) as well as the number of pack years (number of years with an average of one cigarette pack per day). Zero points were assigned to non-smoking, five points to previous smoking, and the maximum of 10 points to current smokers. For converting the number of pack years into scoring points, we used again the classification into quartiles as shown in Supplemental Table 1. The sum of the two scores provided the final smoking score.

Finally, based on the recommendation of the German nutrition society (DGE) we set thresholds for alcohol consumption to create the alcohol score at ten grams per day for females and 20 grams for males. Subjects below or equaling these cut offs scored 0 and those above the thresholds were assigned 5 points.

